# Modifiable lifestyle factors and genetic risk of obesity in Indians

**DOI:** 10.1101/2025.04.18.25326052

**Authors:** Emadeldin Hassanin, Rakesh Kalapala, Nitin Jagtap, Ravikanth Vishnubhotla, Nikhilesh Andhi, Sandeep Mamidi, Sree Vyshnavi, Srikanth Jilla, Mithil Samudrala, P. Shravani Shriya, Kamaldeep Chawla, Carlo Maj, Patrick May, Dheeraj Reddy Bobbili, D. Nageshwar Reddy

**Author notes:** Shared first authorship (Emadeldin Hassanin, Rakesh Kalapala).

## Abstract

This study investigates the potential for individuals of Indian ancestry with an increased polygenic risk score (PRS) for body mass index (BMI) to attenuate their risk of obesity through sustained adherence to a health-promoting lifestyle. For the purposes of this research, a health-promoting lifestyle is defined by routine physical activity, non-smoking or minimal smoking behavior, and the intake of a balanced nutritional diet. We analyzed two independent cohorts: 6,663 Indian participants from the UK Biobank and 91 participants from the Wellytics-Asian Institute of Gastroenterology cohort. Genetic predisposition was quantified using a BMI-PRS, while lifestyle behaviors were combined into a composite score categorized as favorable or unfavorable. Obese individuals exhibited significantly higher PRS values than non-obese counterparts (UKB: P = 5.7 × 10^−27; W-AIG: P = 8.9 × 10^−3). Participants with both a high PRS and an unfavorable lifestyle showed the greatest odds of obesity (UKB: OR = 2.19, P = 3.01 × 10^−9; W-AIG: OR = 16.1, P = 2.99 × 10^−3), whereas those with high genetic risk but favorable lifestyles had reduced odds (UKB: OR = 1.58, P = 3.59 × 10^−3; W-AIG: OR = 3.10, P = 0.129). These findings underscore the potential of lifestyle interventions to attenuate genetically driven obesity risk in this population.

## Introduction

Obesity continues to rise globally and poses a considerable challenge to public health. Although rapidly evolving lifestyles characterized by reduced physical activity and increased intake of calorie-dense foods have accelerated this phenotype^1,2^, twin and family studies, as well as genome-wide association studies (GWAS) underscore the substantial heritability of body mass index^3–5^. The cumulative impact of common genetic variants is often quantified using polygenic risk score (PRS) ^6,7^, providing a means to assess individual susceptibility to obesity^8^.

While genetic predisposition is central to understanding individual variations in obesity susceptibility, equally compelling evidence shows that lifestyle factors - diet, physical activity, and smoking - can modify or even mitigate the phenotypic impact of genetic risk^9–11^. Most research on these interactions has been conducted in Western populations. The Indian population, however, harbors distinct genetic backgrounds and is often susceptible to metabolic complications at lower BMI thresholds, possibly due to distinct genetic architecture and environmental exposures^12–14^. Additionally, rapid cultural and socioeconomic shifts are reshaping traditional Indian diets and physical activity patterns, potentially increasing the prevalence of obesity and related metabolic disorders^15^.

Given these considerations, the present study investigates how lifestyle and genetic risk jointly influence obesity risk in two Indian cohorts: one from the genetically estimated Indian participants from the UK Biobank (UKB) study and another recruited in India through Wellytics and the Asian Institute of Gastroenterology (W-AIG). By employing a PRS for BMI, we aimed to clarify whether adopting a healthy lifestyle can alleviate the impact of high genetic susceptibility on obesity risk. Understanding these patterns could guide the development of targeted strategies for obesity prevention and management in Indian populations and potentially in other high-risk ethnic groups^16^.

## Results

### Participant characteristics

As summarized in Table 1, our final datasets included 6,663 Indian participants from the UKB and 91 from the W-AIG cohort. The ancestry of study participants has been validated by conducting a principal component analysis (PC) on a set of ancestry-informative markers, where it has been observed that Indian participants from both cohorts clustered closely, indicating a high degree of overlap in genetic ancestry between the UKB and W-AIG Indian participants **Supplementary Figure 1**. The UKB sample had a mean age of 53.40 ± 8.41 years, with 67.40% classified as overweight or obese (BMI ≥25 kg/m²). In contrast, the W-AIG cohort was notably younger, with a mean age of 34.23 ± 12.89 years and 53.80% having a BMI ≥25 kg/m².

**Figure 1:**
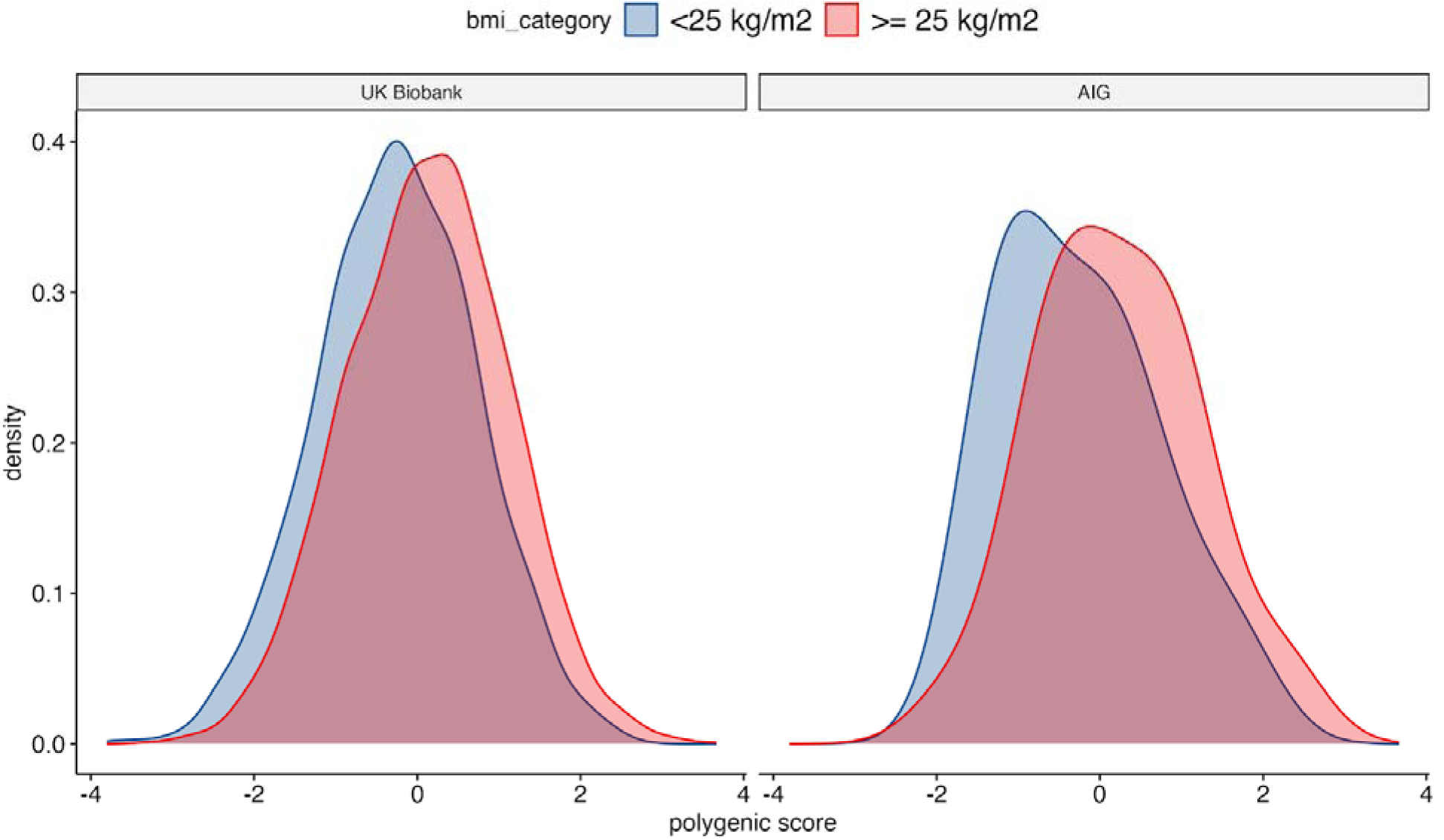
Distribution of aPRS across different obesity groups of Indians participants in UKB and W-AIG cohorts

**Table 1:** Participant characteristics of Indian participants in UK Biobank and W-AIG cohorts.

|  | UKB | W-AIG |
| --- | --- | --- |
| **Participants, N** | 6,663 | 91 |
| **BMI $\geq 25 \text{ kg/m}^2$, N (%)** | 4,490 (67.40) | 49 (53.80) |
| **BMI $< 25 \text{ kg/m}^2$, N (%)** | 1,979 (32.60) | 42 (46.20) |
| **Male, N (%)** | 3,602 (54.10) | 52 (57.14) |
| **Age, mean (SD)** | 53.40 (8.41) | 34.23 (12.89) |

### Polygenic risk score and obesity status

Both cohorts demonstrated a significant difference in aPRS values between obese and non-obese individuals: *P* = 8.9 × 10^−3 in the W-AIG cohort and *P* = 5.7 × 10^−27 in the UKB. This underscores the association between higher polygenic risk and elevated BMI in the Indian population.

### Combined effect of PRS and lifestyle on obesity risk

Logistic regression analyses integrating aPRS category and lifestyle classification are summarized in **Figure 2 and Supplementary table 1**. Across both cohorts, the combination of high PRS and unfavorable lifestyle yielded the highest odds of obesity (UKB: OR = 2.19, P = 3.01 × 10^−9; W-AIG: OR = 16.1, P = 2.99 × 10^−3). Conversely, participants with high PRS who reported a favorable lifestyle had substantially lower ORs (UKB: OR = 1.58, P = 3.59 × 10^−3; W-AIG: OR = 3.10, P = 0.129), though still above unity, indicating a partial attenuation of genetic risk through healthy behaviors.

**Figure 2.**
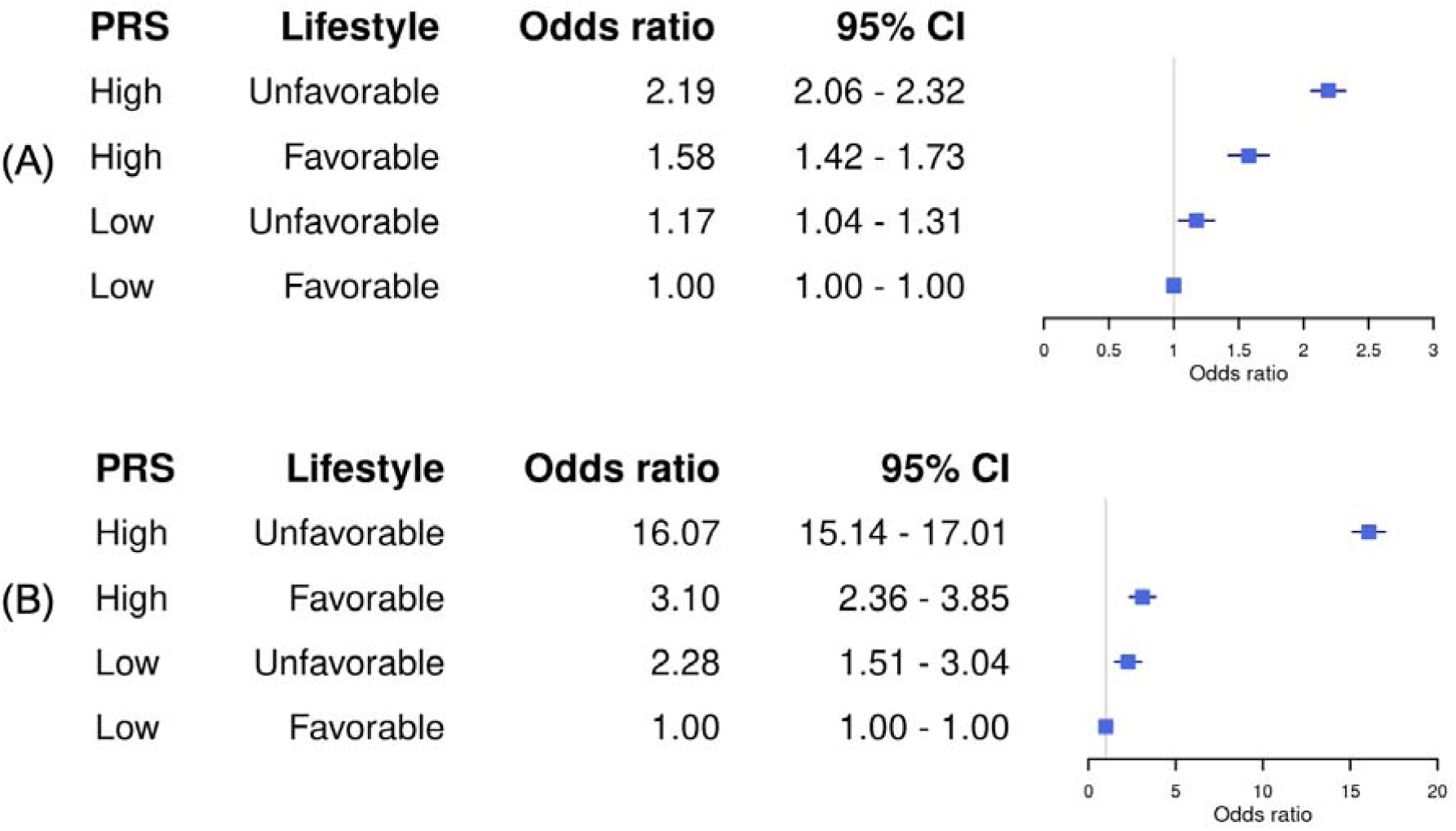
Odds of developing obesity among different genetic and lifestyle categories in the Indian population. (A) UKB; (B) W-AIG cohort.

### Mean BMI across PRS–lifestyle categories

Across both cohorts, participants with high genetic risk and unfavorable lifestyles had the highest mean BMI values. Notably, high aPRS individuals with favorable lifestyles had lower mean BMIs than those with unfavorable lifestyles, suggesting that healthy lifestyle behaviors can mitigate the impact of genetic predisposition on BMI (**Figure 3 and Supplementary table 2**).

**Figure 3:**
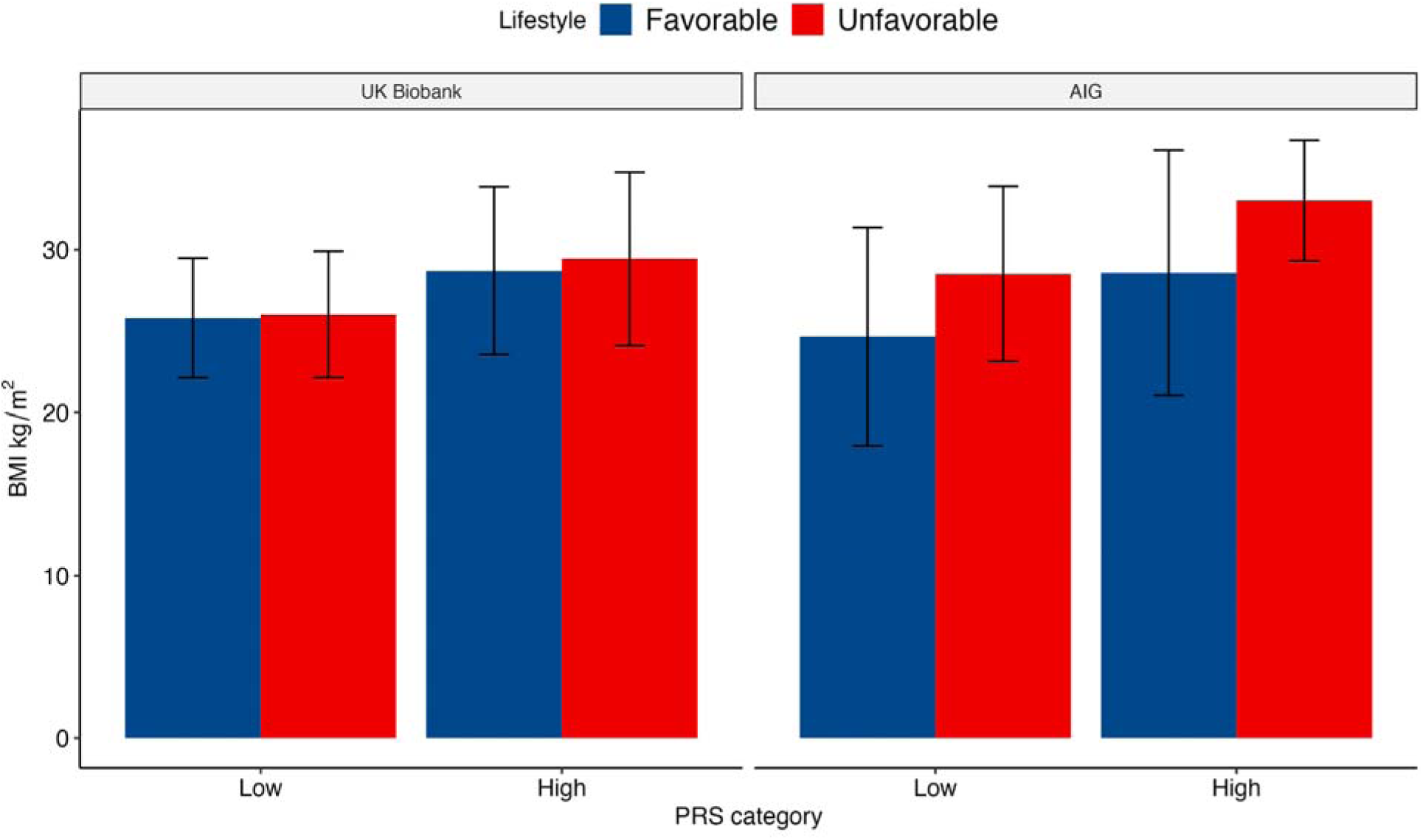
Mean BMI across PRS - lifestyle categories of Indian participants in the UKB and W-AIG cohorts

## Discussion

The risk of obesity among Indian adults can be significantly affected by modifiable lifestyle factors, which can potentially offset the genetic predisposition for obesity^17^. Following a few studies in the European population^9,11,18^, our research using two Indian cohorts strengthens the understanding that while a high PRS substantially increases the risk of obesity, this risk can be mitigated by environmental and behavioral factors that can be modified by individuals and public health systems.

The W-AIG cohort, with a mean age in the mid-30s, displayed a substantial OR of 16.1 for high PRS and unfavorable lifestyle Supplementary table 1. This suggests that lifestyle choices may have a greater impact on individuals with obesity-linked alleles at younger ages. However, it should be noted that the relatively small sample size of the W-AIG cohort might have also contributed to the large effect size observed. In contrast, the UKB participants, who are older on average, showed smaller effect sizes for similar PRS and lifestyle patterns, although they were still significant. This difference may be attributed to accumulated metabolic changes or decreased plasticity over time, which could moderate the gene-environment interaction in older adults. Despite this, the overall pattern of interaction remains consistent across age groups.

A similar divergence emerges in the mean BMI analysis. W-AIG participants with high PRS and unfavorable lifestyle exhibit a large gap relative to their counterparts with a favorable lifestyle (∼4.4 kg/m² difference), whereas the UKB cohort shows a narrower gap (∼0.8 kg/m²) Supplementary table 2. This difference could reflect more extreme lifestyle behaviors in certain W-AIG subgroups or a cultural context that amplifies the deleterious effects of poor diet and limited physical activity. Alternatively, the small sample size in W-AIG could inflate effect estimates, underscoring the need for replication in larger Indian cohorts. Nonetheless, the consistency of the trends underscores the central finding: a synergy exists between genetic predisposition and lifestyle that can drive obesity risk upward or, conversely, offer protection.

Notably, participants with high PRS but favorable lifestyles in both cohorts had reduced odds of obesity compared to their counterparts with unfavorable lifestyles, pointing to a partial but meaningful mitigation of genetic risk. These findings strongly advocate for targeted health interventions focusing on smoking, regular exercise, and a diet less reliant on refined carbohydrates and saturated fats - behaviors that appear particularly important for individuals at high genetic risk. Public health initiatives that facilitate early identification of such high-risk individuals may yield efficient strategies to curb obesity-related morbidity^19,20^. Equally important, ongoing research should investigate how finer gradations of diet (e.g., precise macronutrient composition) and exercise (e.g., intensity, duration) interact with specific genetic sub-profiles to yield even more tailored interventions.

Beyond genetics and lifestyle, socio-cultural and economic factors may also shape individuals’ diets, access to exercise, and overall health-seeking behaviors. Indians living in the UK might have a different lifestyle compared to those residing in India^21,22^, influencing how much an “unfavorable lifestyle” truly differs between these settings. Further, a cross-sectional study can only capture a snapshot; prospective or longitudinal studies would highlight whether consistent adherence to favorable lifestyle behaviors over years can attenuate genetic risk in a more durable manner. Evidence like this could influence lifestyle changes that put a focus on early behavioral interventions - possibly during childhood or young adulthood - for those with a higher genetic predisposition.

A key strength of this study is the replication of findings in two independent cohorts, one from a Western context (UKB) and another recruited directly in India (W-AIG). This design enhances the external validity of our findings and confirms that the gene–lifestyle interaction is not exclusive to a single environment. Moreover, using a PRS consolidates the polygenic underpinnings of obesity, offering a broad perspective on genetic liability.

The current research, however, has limitations. First, our lifestyle metrics were self-reported and dichotomized, which could obscure nuanced differences in diet and physical activity. Additionally, the relatively small W-AIG sample size may inflate effect size estimates and limit statistical power. Finally, the cross-sectional design of our study prevents determination of causality or the long-term sustainability of lifestyle changes. Future research should utilize larger, longitudinal studies with refined phenotyping to address these limitations, which would allow for stronger causal conclusions and more precise public health recommendations.

In summary, to our knowledge, our study shows for the first time that lifestyle choices play a key role in obesity risk among Indian individuals with high genetic risk. Even with a strong genetic predisposition, obesity is not inevitable. We found that those with high genetic risk and an unhealthy lifestyle had higher odds of obesity, while those with healthier behaviors had lower odds. This supports the need for public health actions promoting better diets, regular exercise, and tobacco avoidance. Future studies should explore more detailed lifestyle factors, long-term trends, and potential differences based on sex or region. Ultimately, helping at-risk groups adopt healthier choices can be a powerful tool in reducing obesity and related health problems.

## Methods

### Study design and participants

We conducted a dual-cohort study using data from the UK Biobank (UKB) and Wellytics-Asian Institute of Gastroenterology (W-AIG) cohorts. The UKB data included 6,663 participants of genetically-estimated Indian ethnicity confirmed via principal component analysis of genetic data. The W-AIG study recruited 91 Indian participants from diverse regions across the country. All participants in both cohorts provided written informed consent. Ethical approvals were obtained from the NHS National Research Ethics Service (reference 11/NW/0382) for UKB and the Institutional Ethics Committee of the Asian Institute of Gastroenterology (approval number AIG/IEC-CT63/06.2022-03) for W-AIG.

### UK Biobank cohort

The UKB is a large, prospective study of over 500,000 individuals aged 37 to 73 from England, Scotland, and Wales^16^. We extracted data on individuals of Indian ancestry, wherein BMI was calculated using measured height and weight. Following World Health Organization (WHO) guidelines adapted for Asian populations^23^, obesity was defined as a BMI ≥25 kg/m². Lifestyle information was self-reported, covering dietary patterns (e.g., fruit and vegetable intake), physical activity (frequency and intensity), and smoking status^18^. Imputation and QC of the UKB data have been described elsewhere^16^.

### Wellytics–Asian Institute of Gastroenterology (W-AIG) cohort

**Genotyping data and QC:** The W-AIG cohort comprised 91 adults who underwent saliva-based genotyping using Genome-Wide SNP Array V3 (GSA V3). We excluded samples with more than 1% missing genotypes and variants exceeding 1% missingness. Only biallelic SNPs (A, C, G, T) were retained. Any samples where there is a discrepancy between the predicted gender and the reported gender were excluded. Subsequently we merged our cleaned dataset with the UKB dataset for population stratification. We performed imputation by harmonizing variant naming and strand orientation by employing the HRC-1000G-check-bim script, filtering out discordant positions before phasing with Eagle (using 1000 Genomes Phase 3 as a reference) and imputing via Beagle^24,25^. Post-imputation, variants with low-quality dosage scores (DR2 < 0.3) were excluded, ensuring a high-confidence dataset.

**Clinical data:** Participants were categorized as healthy (BMI <25 kg/m²) or overweight/obese (BMI ≥25 kg/m²). A standardized questionnaire collected details on participants’ socioeconomic status, diet, physical activity, psychosocial factors, medications, and more. This approach yielded a comprehensive picture of lifestyle behaviors pertinent to obesity risk.

**Principal component analysis:** To assess population structure, we first extracted HapMap SNPs from both the W-AIG and UKB datasets. The two datasets were then merged based on the common overlapping variants to ensure consistency in variant representation. The merged dataset was then projected onto the principal component space of the 1000 Genomes Project to predict the ethnicity using *bigsnpr* R package^26^. This enabled us to capture global ancestry patterns and account for potential population stratification in downstream analyses.

### Polygenic risk score calculation

A PRS for BMI was computed through the Wellytics platform using SNPs known to be associated with obesity risk, with effect sizes drawn from the Polygenic Catalog Score^27^ (PGS; PGS000027). Ambiguous variants were excluded, and allele flipping was performed where necessary. To adjust for ancestral background, we fitted a linear regression of the raw PRS on the first four principal components. The predicted component was subtracted from the raw PRS, and the residual was standardized to generate an adjusted PRS (aPRS). Each cohort’s aPRS distribution was then split into high vs. low groups (using a median split) for further analysis^28,29^.

### Lifestyle score

A lifestyle score was calculated based on three modifiable lifestyle components: smoking, physical activity, and diet^30,31^. Favorable lifestyle factors were defined as following:

1. Smoking: No smoking or moderate consumption of less than 2 cigarettes a day.
2. Physical activity: In the W-AIG cohort, at least 5 days of physical activity, and in the UKB cohort at least 5 days of a combination of moderate physical activity and vigorous activity.
3. Healthy diet: Based on consumptions of fruits of fruits, vegetables, fish, whole grains, refined grains, processed and unprocessed meat and defined as follows:

a. Total fruit intake: 1 to 3 pieces or servings a week.
b. Total Vegetable intake: 1 to 3 pieces or servings a week.
c. Total Fish intake 1 to 3 servings a week.
d. Total whole grains intake: 1 to 3 servings a week.
e. Total refined grains intake: <= 1 servings a week.
f. Total meat intake: <= 1 servings a week.

The healthy diet score was defined as an individual meeting at least four of the healthy food items listed above.

Each of the three components was assigned a score of 0 or 1, with 1 indicating the healthiest behavior. The lifestyle score was calculated by summing the scores for each component, with a maximum score of 3 indicating the healthiest lifestyle. The lifestyle score was dichotomized into:

- Favorable: Scored ≥ 2 of the healthy lifestyle factors.
- Unfavorable: Scored < 2 of the healthy lifestyle factors.

### Statistical analysis

Participants were stratified into four groups based on their aPRS (low or high) and lifestyle score (favorable or unfavorable):

1. Low aPRS, Favorable Lifestyle (reference group)
2. Low aPRS, Unfavorable Lifestyle
3. High aPRS, Favorable Lifestyle
4. High aPRS, Unfavorable Lifestyle

Logistic regression models were used to assess the association of aPRS and lifestyle with obesity risk, as well as their combined effect. Odds ratios (ORs) and standard errors were calculated to estimate the effect sizes, using the “Low aPRS, Favorable Lifestyle” group as the reference. Models were adjusted for age, sex, and the first four ancestry PCs to control for potential confounding factors.

To test for a statistical interaction between lifestyle and genetic risk in relation to obesity, an interaction term between aPRS and lifestyle score was included in the regression models. Statistical significance was set at p < 0.05. All statistical analyses were performed using R statistical software (version 4.2.2).

## Declarations

### Consent for publication

Not applicable.

### Data Availability

Data used to prepare this article were obtained from UKB. Restrictions apply to the availability of these data for UKB, which were used under license for the current study (Project ID: 52446).

### Authors’ contributions

(1) Research Project: A. Conception, B. Organization, C. Execution; (2) Statistical Analysis: A. Design, B. Execution; (3) Data: A. acquisition B. Curation (4) Manuscript Preparation: A. Writing of the First Draft, B. Review and Critique.

E.H: 1A, 1B, 1C, 2A, 2B, 3B, 4A, 4B

R.K: 1A, 1B, 1C, 2A, 3A, 4A, 4B

N.J: 1A, 1B, 1C, 3A, 4A, 4B

R.V: 1A, 1B, 1C, 3A, 4A, 4B

N.A: 1A, 1B, 1C, 3A, 3B, 4A, 4B

S.M: 1B, 1C, 4A, 4B

S.V: 1B, 1C, 4A, 4B

P.S.S: 3A, 3B, 4A

K.C: 4B

C.M: 4B

P.M: 4B

D.R.B: 1A, 1B, 1C, 2A, 2B, 3A, 3B, 4A, 4B

D.N.R: 1A, 1B, 1C, 2A, 2B, 4A, 4B

## Acknowledgements

Data used to prepare this article were obtained from the UKB. Ethics approval for the UK Biobank (UKB) study was obtained from the Northwest Multicentre for Research Ethics Committee (MREC). The UKB ethics statement is available at https://www.ukbiobank.ac.uk/learn-more-about-uk-biobank/about-us/ethics. All UKB participants provided informed consent at recruitment. Similarly, all W-AIG participants provided written informed consent, and the study was approved by the Institutional Ethics Committee of the Asian Institute of Gastroenterology (approval number AIG/IEC-CT63/06.2022-03). Parts of the computational analysis were done on the High-Performance Computing cluster of the University of Luxembourg (https://hpc.uni.lu/).

